# Study of an age-based Covid-19 outbreak model and the effect of demographic structure of a state on infectious disease dynamics

**DOI:** 10.1101/2022.12.28.22284021

**Authors:** Ayanava Basak, Tanmay Sanyal

## Abstract

In this paper, our objective was to investigate whether the Covid-19 pandemic disease is more likely to break out in some specific age group or not. We also intended to know whether some specific demographic parameters like birth rate, death rate controls the spreading of the disease. Our investigation showed that the post reproductive population group is more prone to the disease for the countries having population pyramid of stationary or contracting type where as for the countries with expanding population pyramid, the pre reproductive population is more likely to be attacked by the disease. We also found the domains of values of the demographic parameters that result different dynamic phenomena. Further we tried to know whether a country’s population pyramid has an effect in spreading the disease. Our experiment showed that for countries having expanding population pyramid, the total number of cases is expected to be comparatively low whereas for countries having contracting population pyramid, the total number of cases is expected to be comparatively high.

## Introduction

The nature of the dynamics of evolution of Covid-19 pandemic is different in different countries (Coronavirus graphs: worldwide cases and deaths - worldometer). A lot of reasons might be present behind this. Researchers have found the probable causes can be the diversity in the population distribution (Wong and Li, 2020), the diversity in the climate (Brassey et al., 2020; Lin et al., 2006; Chan et al., 2011), latitude and longitude (Burra et al., 2021; Salman et al., 2020) or Non-Pharmaceutical Interventions (NPI) (Flaxman et al., 2020). In this article we tried to investigate the dynamics of Covid-19 pandemic in several countries, belonging to different population pyramid. Population pyramids are classified into three different types- (1) Expanding (2) Stationary (3) Contracting (Saroha, 2018; World demographics 2020 (Population, age, sex, trends) - worldometer). Every country belongs to a particular population pyramid based on their age and sex structure (Saroha, 2018). Also, the gradual change of fertility, natality and mortality of different age groups control the demography of a state. Critical changes of such controlling factors take a nation from one population pyramid to another which is known as Demographic Transition (Saroha, 2018). Also, it is known that some infectious diseases have the tendency to affect a particular age group of population. For example, AIDS is a disease for which the incidence is high in the age group from 20 to 45 (Martcheva, 2015). Here we focused on two principal facts: firstly, how the birth and death rates of different age structures of a population influence the outbreak of Covid-19 pandemic and secondly, whether the Covid-19 is more prone to attack some specific age group of a population.

We introduced an age-based infectious disease evolution model. Then we chose three countries having three different categories of population pyramid and obtained the time series of the model for particular values of the involved parameters for better understanding of the disease dynamics. We checked how the initial demographic state i.e., the initial population of different age groups, influence the total number of cases. After that we analyzed the effect of different demographic parameters on the total number of cases in different age groups by one parameter, two parameter and three parameter bifurcation diagrams. We found that like AIDS, Covid-19 is also more likely to break out in particular age group depending on the demographic state of the country. The obtained results have been discussed.

This paper contains seven sections. Section I explains the methods and techniques that have been adopted. In Section II, the formation of the model is done along with the required assumptions. The parameters were addressed there as well and the time series was also included in that section. In Section III, the initial value sensitivity was discussed. Section IV and V contain one and two parameter bifurcation respectively and in Section VI there is the notion of three parameter bifurcation, which to our knowledge is not discussed anywhere else. We discussed the dependence of total number of cases on initial population in Section VII.

### Section I: Methods and Techniques

First of all, we solved the age based infectious disease model by numerical simulation using Runge-Kutta method. Thus, the time series was obtained and to determine the cumulative number of cases from the time series, obtained, Trapezoidal rule was applied. One parameter, two parameter and three parameter bifurcations were performed with respect to three important parameters to determine the change of the dynamics. Initial value sensitivity was also done to analyze the system. All the figures, graphs and diagrams were obtained using Python software.

### Section II: Formation of Mathematical Model

#### A. Assumptions

1. We divide the total population into three different age groups- (a) Pre-Reproductive Population i.e., the population of age under 18 years (b) Reproductive Population i.e., the population of age between 19 to 45 years. (c) Post Reproductive Population i.e., the population of age above 45 years.
2. Each age group is further subdivided into two distinct classes-Susceptible and Infected Class. Thus, the variables can be described as follows: S_1_ = Susceptible Pre-Reproductive Class I_1_ = Infected Pre-Reproductive Class S_2_ = Susceptible Reproductive Class I_2_ = Infected Reproductive Class S_3_ = Susceptible Post Reproductive Class I_3_ = Infected Post Reproductive Class
3. The time is considered in days.
4. We assume that both the susceptible and infected reproductive population gives birth to new population at a rate, proportional to the size of reproductive population and the new born population is immediately added to the susceptible pre reproductive group.
5. The Covid-19 disease has been found not to be transmitted vertically (Fan et al., 2021) i.e., the new born babies are not infected directly from the placenta of infected mother. So, both the susceptible and infected population gives birth to susceptible population only.
6. It is assumed that the susceptible and the infected population are mixed homogenously and the disease spreads due to body contact following the Mass Action Law with frequency dependent transmission.
7. Suppose the dynamics of the disease incidence follows the SIS model i.e., the infected population recovers and leaves the infected class at a rate proportional to the size of infected population and again joins the susceptible group. Thus, each infected individual spends 1/μ time in the infectious class.
8. Let the pre reproductive, reproductive and post reproductive class die at rates proportional to their sizes respectively.
9. We also assume that the infected population has a higher mortality than the susceptible population.
10. The susceptible pre reproductive and reproductive groups leave their classes at a rate proportional to their respective size. Thus 1/g_1_ and 1/g_2_ are the time (in days) that each susceptible pre reproductive and reproductive individual spends in their respective groups.
11. Since the time that an individual spends in infectious class is very small, we consider that the infected populations do not grow and transmit into higher age group.
12. We consider that the susceptible reproductive and post reproductive groups are vaccinated at a rate, proportional to their sizes respectively.
13. We do not consider immigration and emigration into account.
14. Further we assume that the disease does not affect the reproductive organs i.e., infected individuals are as reproductive as the susceptible.
15. It is also assumed that the susceptible individuals when leave their corresponding age group, join the susceptible class of next age group.
16. Finally, we consider the vaccine to be 100% effective.

Based on the assumptions we consider the following model-

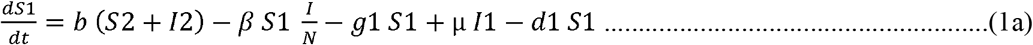

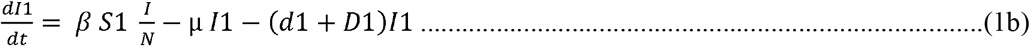

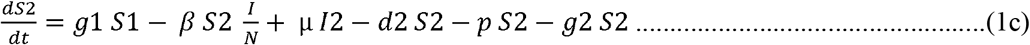

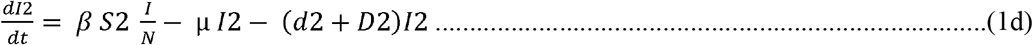

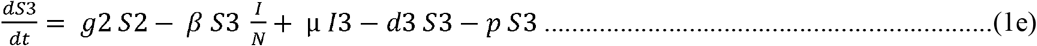

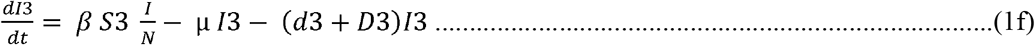

Here I = I1+I2+I3 and N = S1+S2+S3+I1+I2+I3.

We draw a rough schematic diagram of the model formation-

#### B. Introduction to parameters

**Table 1:** Description of different biological parameters, involved in the model.

| Parameters | Significance |
| --- | --- |
| b | Natality or the number of new born children per 1000 people in one day |
| $\beta$ | Disease transmission coefficient |
| $1/g_1$ | Time spent by each pre reproductive individual in that class |
| $1/g_2$ | Time spent by each reproductive individual in that class |
| $1/\mu$ | Time spent by each infected individual in that class |
| $d_1$ | Natural mortality of pre reproductive population in absence of disease |
| $d_2$ | Natural mortality of reproductive population in absence of disease |
| $d_3$ | Natural mortality of post reproductive population in absence of disease |
| $D_1$ | Disease induced mortality of pre reproductive population |
| $D_2$ | Disease induced mortality of reproductive population |
| $D_3$ | Disease induced mortality of post reproductive population |
| $p$ | Rate of vaccination |

#### C. Values of parameters

If 20 new individuals are born from a population of 1000 people in a year, we will take birth rate as (20/365) ≈ 0.0548 per day. From (Birth rate, crude (Per 1,000 people) | Data) we see that the number of new born individuals vary from 6 to 46. Usually, birth rates ranging from 10 to 20 births per 1,000 are considered low, while rates from 40 to 50 births per 1,000 are considered high (Birth rate. In: Wikipedia. 2022). So, the birth rate can vary from 0.016 to 0.126 per day. We will vary it in the range 0.0 to 0.15 per day. In a similar fashion we also vary d_1_ and d_2_ from 0.0 to 0.15 per day. Using the notion, introduced in (Biswas et al., 2020) d_3_ can also be determined easily. Suppose 75 is the average life span of a country then the death rate of the people of age more than 45, is- 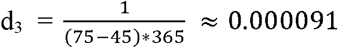 per day. We assume D_1_=D_2_=D_3_ and choose the value as in (Biswas et al., 2020).

From the definition, we calculate g_1_ and g_2_ to be 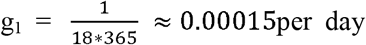 and 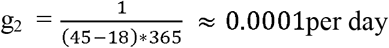.

Now we calculate the rate of vaccination. Though it is different in different nations, in this paper we will consider it same for all countries. We determine the rate of vaccination of India. The number of fully vaccinated persons in India is 30,45,91,540 (Cowin dashboard) as of 24^th^ October,2021 from 13^th^ February,2021 and the total population of India as of 24^th^ October,2021 is 1,39,77,53,851 (India population (2022) - worldometer). So, the rate of vaccination is, 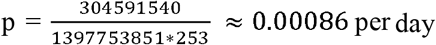.

#### D. Numerical Simulation and Time Series

We choose three countries belonging to three distinct population pyramids (Saroha, 2018). The choice is completely arbitrary. We find the percentage of their population belonging to different age groups from (Population Pyramids of the World from 1950 to 2100. PopulationPyramid.net). Next, we consider the initial population of each country to be 1000 and then determine the initial number of individuals belonging to each age group. Finally, we find the values of b, d_1_ and d_2_ for each country from (Birth rate, crude (Per 1,000 people) | Data; Population Pyramids of the World from 1950 to 2100. PopulationPyramid.net;) and take the values of other parameters as in Table 2. Thus, we obtain the time series for these countries with the time varying from 0 to 500 days, shown in Figure 2. The descriptions of the countries are given in TABLE-3. It is also assumed that 1% of pre reproductive population is infected, 2% of reproductive population and 3% of post reproductive population is infected initially.

**Table 2:**
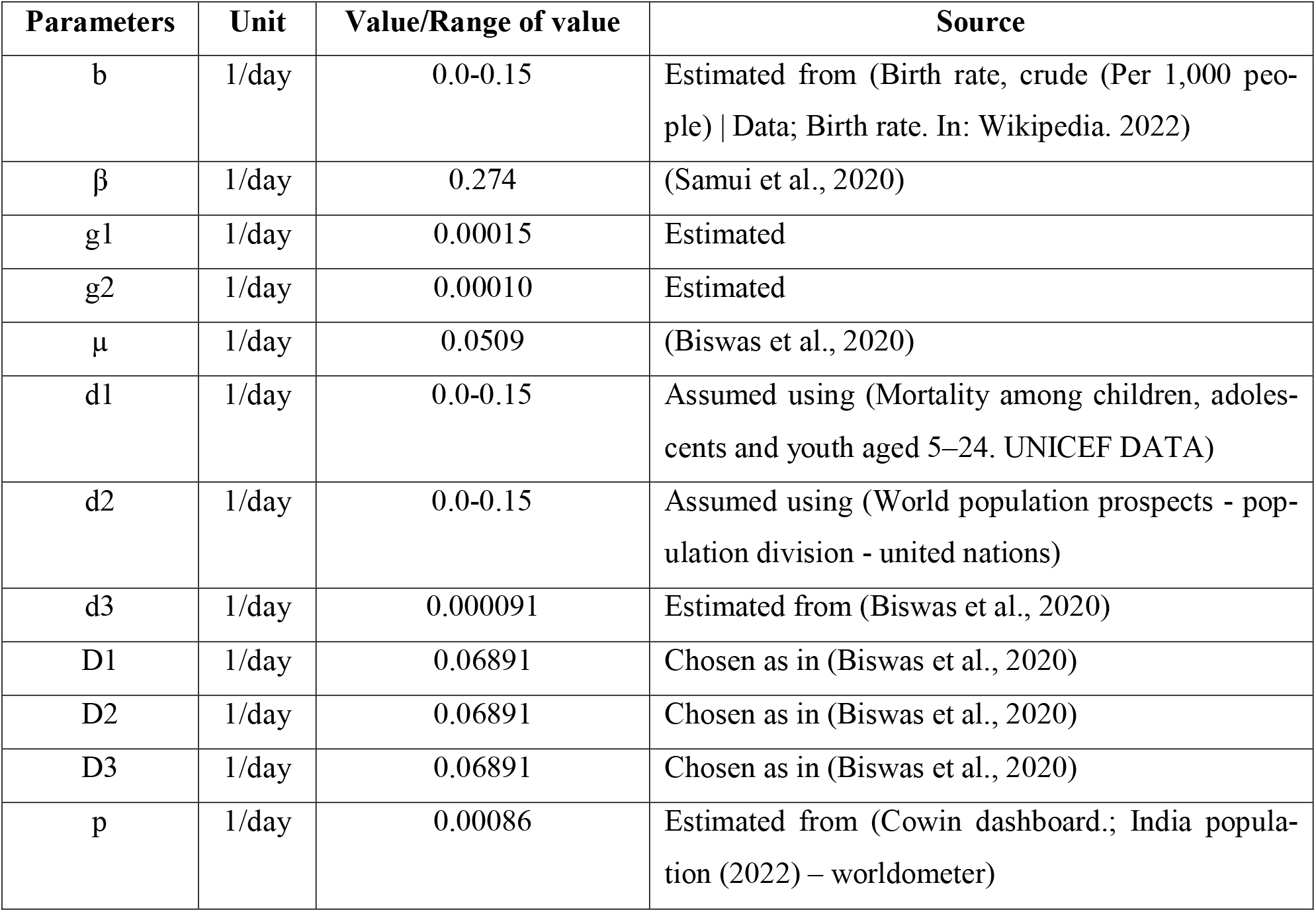
Determination of values of different parameters.

**Table 3:**
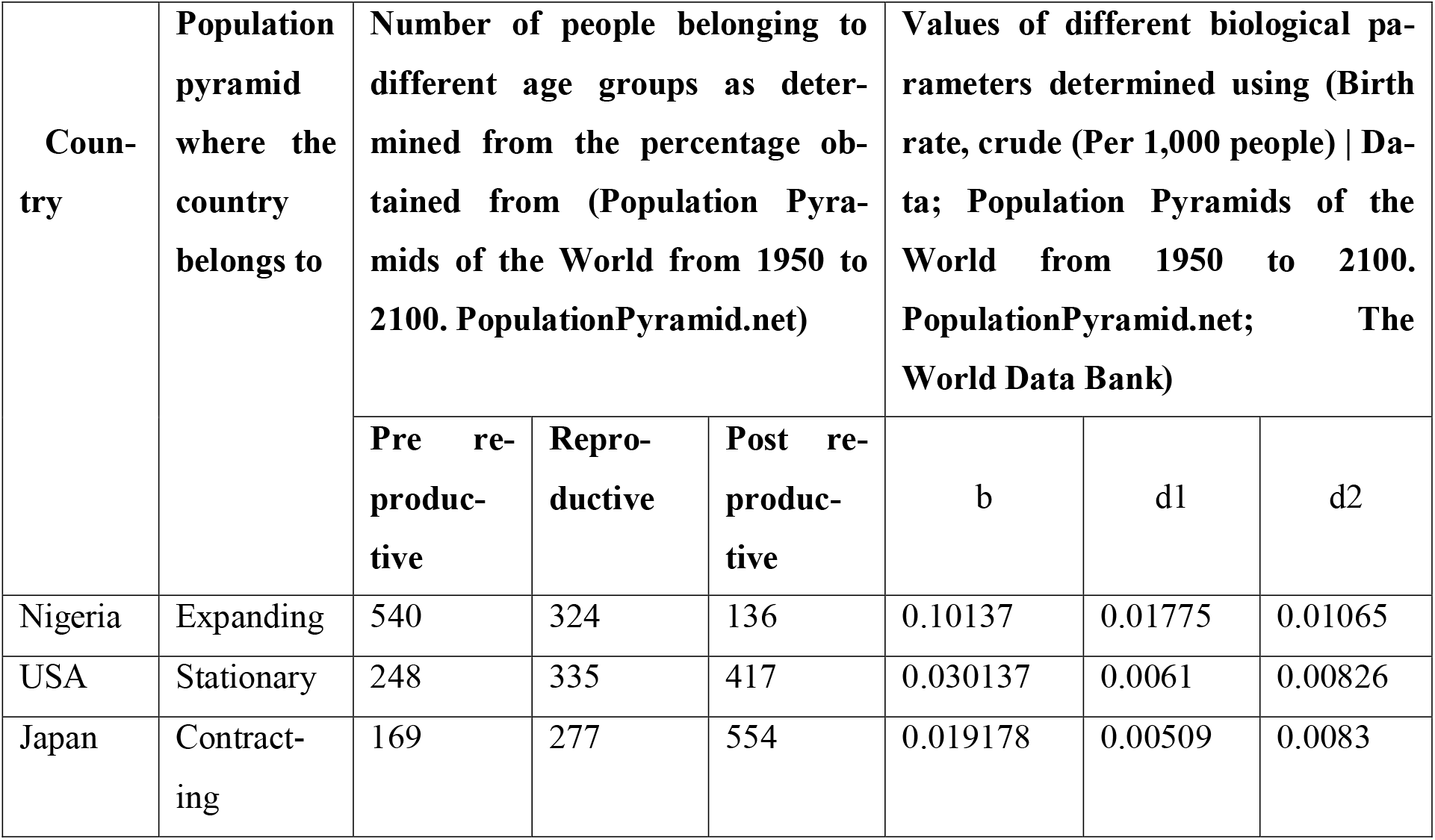
Determination of the values of b, d1 and d2 for different countries.

Now we plot the time series of these three countries belonging to three different population pyramids in Figure 2(a), 2(b) and 2(c). In all the figures, solid blue line shows susceptible pre reproductive population and similarly solid red, black, green, purple, olive-green lines represent susceptible reproductive, susceptible post reproductive, infected pre reproductive, infected reproductive and infected post reproductive population respectively. The black broken line gives the total infected population. The area under each curve gives the total number of individuals of each class-which can be calculated easily using any simple numerical method. The results obtained from this numerical simulation, are given in the observation part.

**Figure 1:**
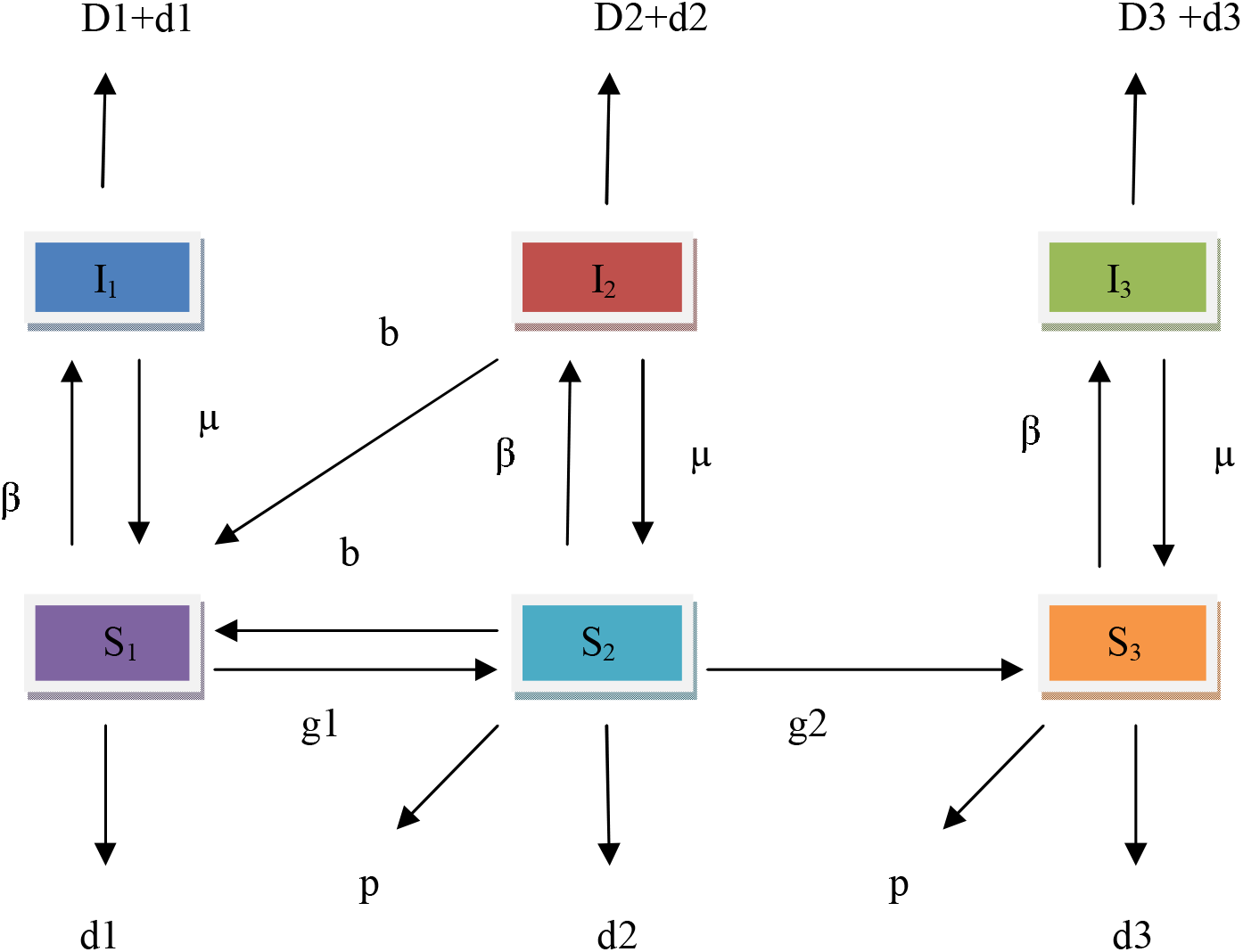
Schematic diagram of age-based infection model.

**Figure 2(a).**
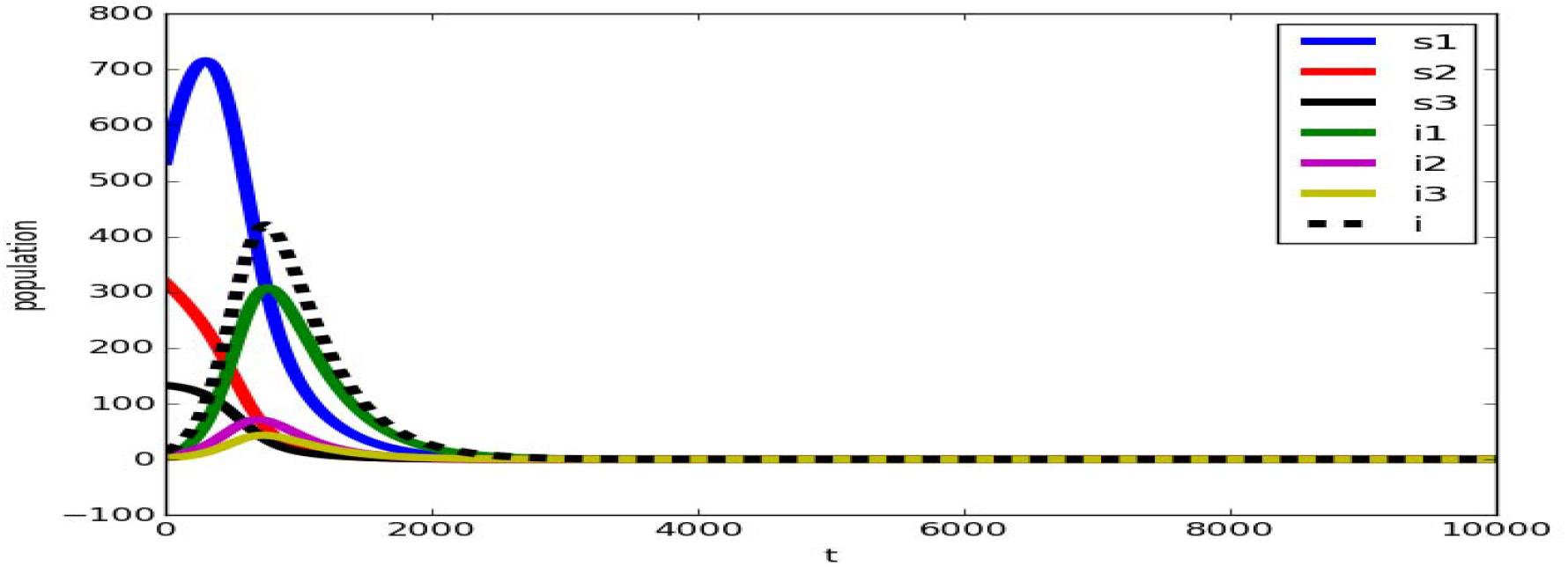

**Figure 2(b).**
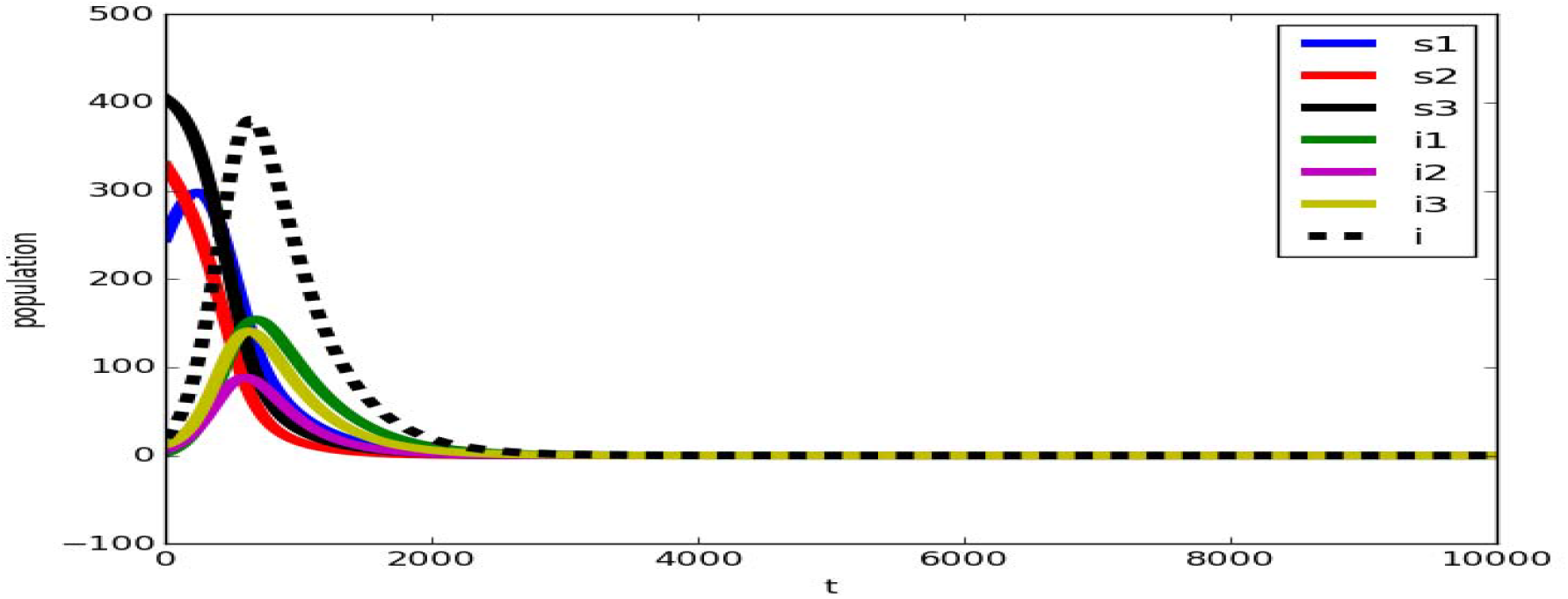

**Figure 2(c).**
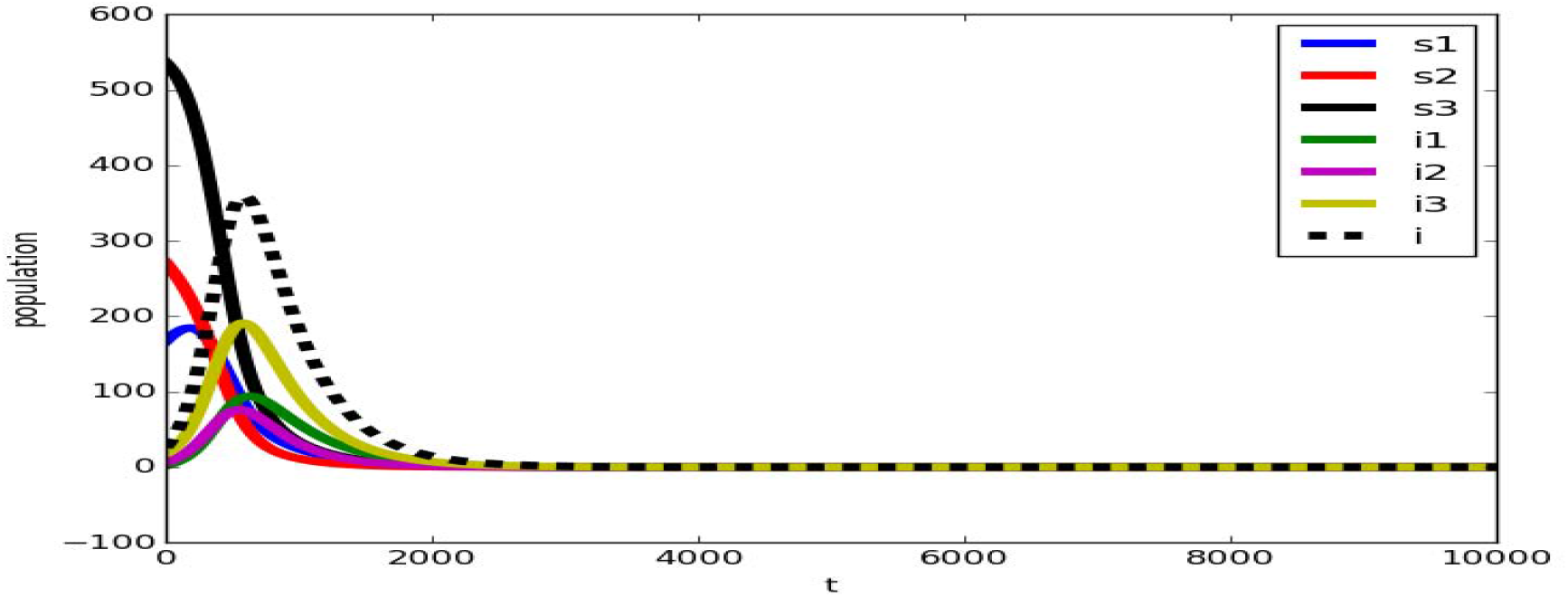

#### E. Results and Observations

First of all, let us show the results obtained from this numerical simulation.

We see that, here the number of infected individuals is highest when the country has a population pyramid of expanding type. And the number of cases is lowest when the country belongs to a population pyramid of contracting type.

Further we observe that the share of infected population, occupied by each age group, is dependent on the type of that country’s population pyramid. Pre reproductive group occupies a high percentage (73.67%) of total infected people for Nigeria. This is quite obvious because a country with expanding population pyramid has a huge portion of pre reproductive people which is not vaccinated according to our assumption. This result also suggests that in those countries having expanding population pyramid, special precautions should be taken for pre reproductive population.

More than half (52.79%) of total cases belong to post reproductive population in Japan. So, in spite of vaccination, for the countries having population pyramid of contracting type, the post reproductive group is at high risk. Reproductive group is not at high threat in all kind of population pyramid. The vaccinated class is also prone to be infected more in the countries whose population pyramid is not expanding type.

### Section III: Initial Value Sensitivity

Let TI_1_ = total number of infected pre reproductive population

TI_2_ = total number of infected reproductive population

TI_3_ = total number of infected post reproductive population.

Now six cases may arise: Case-I when TI_3_ TI_2_ TI_1_(denoted by c1); Case-II when TI_2_ TI_3_ TI_1_(denoted by c2); Case-III when TI_2_ TI_1_ TI_3_(denoted by c3); Case-IV when TI_3_ TI_1_ TI_2_(denoted by c4); Case-V when TI_1_ TI_3_ TI_2_(denoted by c5); Case-VI when TI_1_ TI_2_ TI_3_(denoted by c6). From previous observation, we see that these six cases arise when the initial value and the parameters are varied. In this section we vary the initial populations of all three age-based groups from 0 to 1000 keeping the total population constant at 1000 and plot those values with different colors indicating the above six cases. We choose b = 0.053 per day, d_1_ = 0.049 per day and d_2_ = 0.019 per day and other parameters same as Table 2.

It is found (Figure 3(a)) that the plane x + y +z =1000 is divided into six different regions of different colors. The blue region suggests c1; sea green region suggests c2; purple color gives c3; red for c4; olive green denotes c5 and the green region gives c6. We also find the approximate percentage of area of each region by calculating the percentage of points which represent each condition above and represent them with a pie diagram shown in Figure 3(b).

**Figure 3(a).**
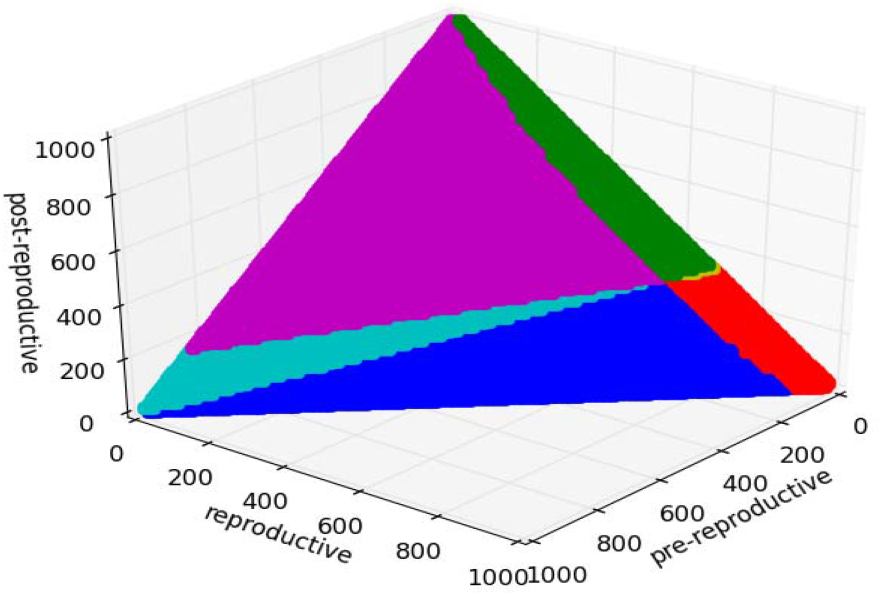

**Figure 3(b).**
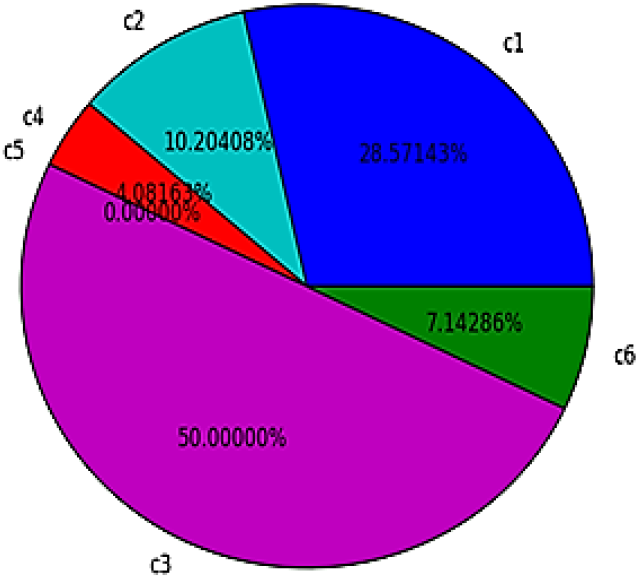

### Section IV: One Parameter Bifurcation

We have chosen b, d_1_ and d_2_ as the bifurcating parameters for their utmost importance. Here we increase any one of these parameters from 0.0 to 0.15; keeping all the other parameters fixed and plot the number of infected pre reproductive, reproductive post reproductive and total number of cases for all the three countries in Table 3. Figure 4(a), 4(b), 4(c) are the bifurcation diagrams of Nigeria with respect to b, d_1_ and d_2_ respectively. Similarly Figure 4(d), 4(e), 4(f) are the bifurcation diagrams of USA with respect to b, d_1_ and d_2_ respectively; Figure 4(g), 4(h), 4(i) are the bifurcation diagrams of Japan with respect to b, d_1_ and d_2_ respectively. Solid green, purple and olive-green curve represents pre reproductive, reproductive and post reproductive population respectively. Black dotted line denotes the total number of cases.

**Figure 4(a).**
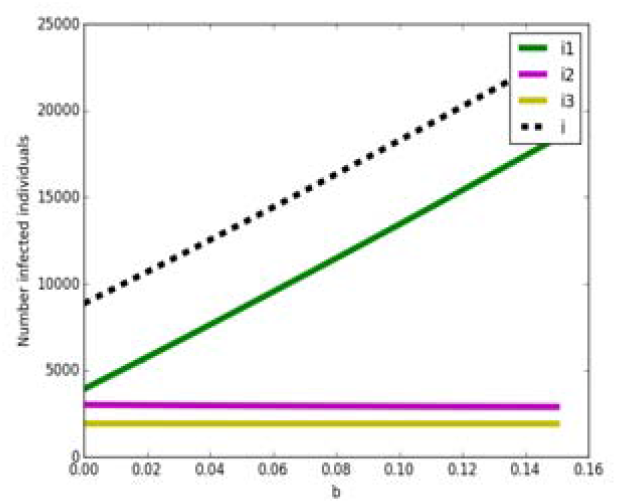

**Figure 4(b).**
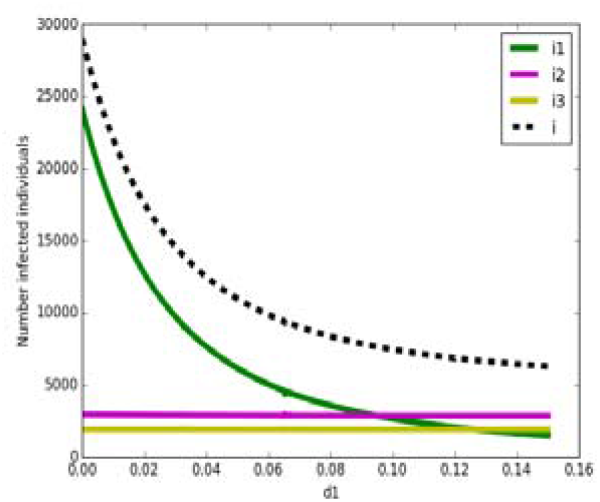

**Figure 4(c).**
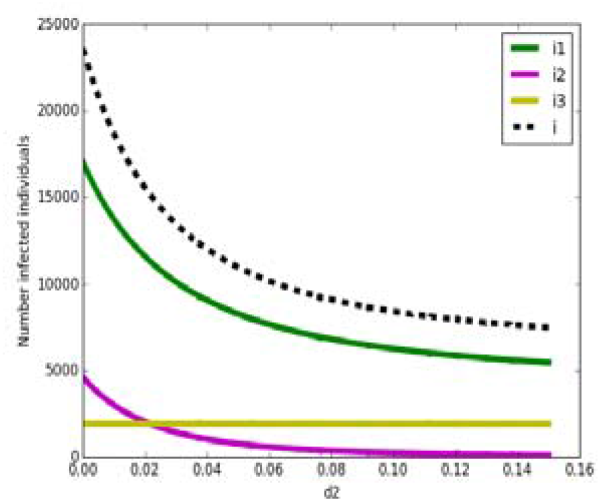

**Figure 4(d).**
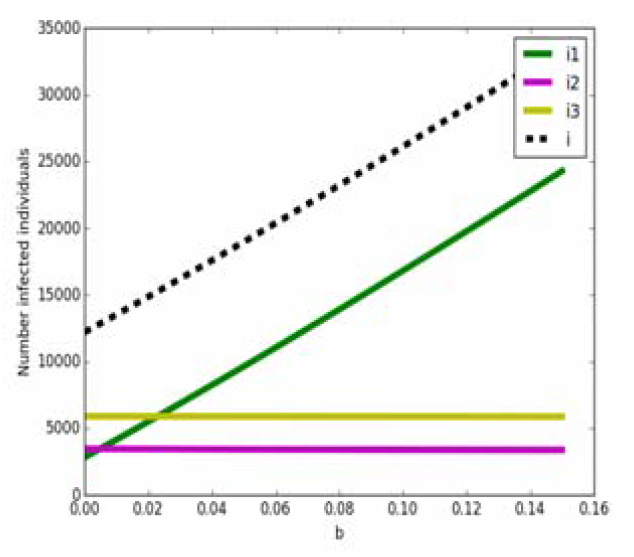

**Figure 4(e).**
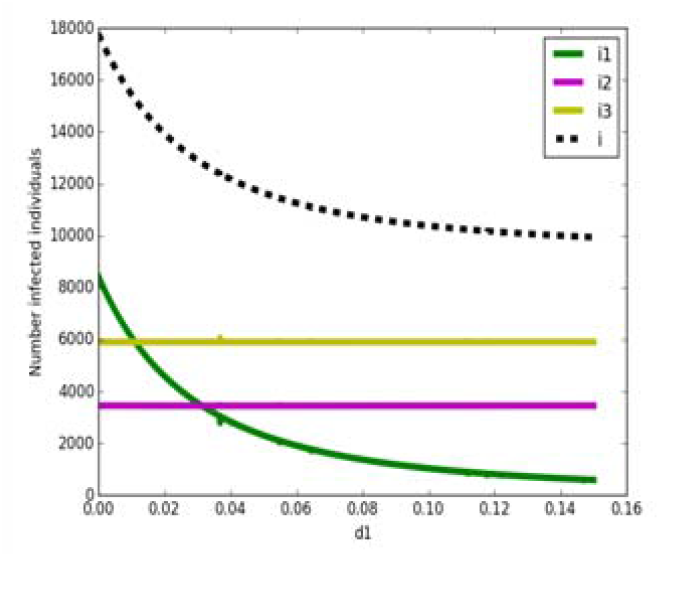

**Figure 4(f).**
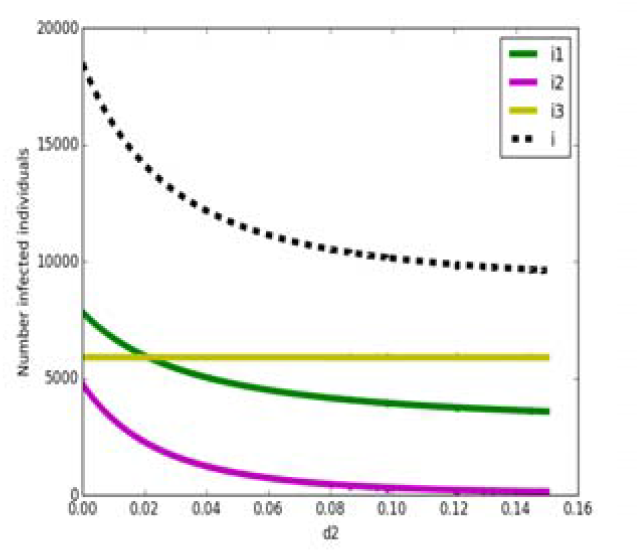

**Figure 4(g).**
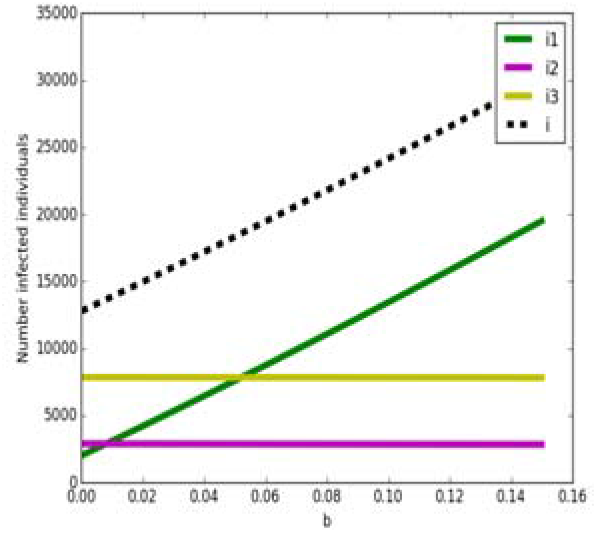

**Figure 4(h).**
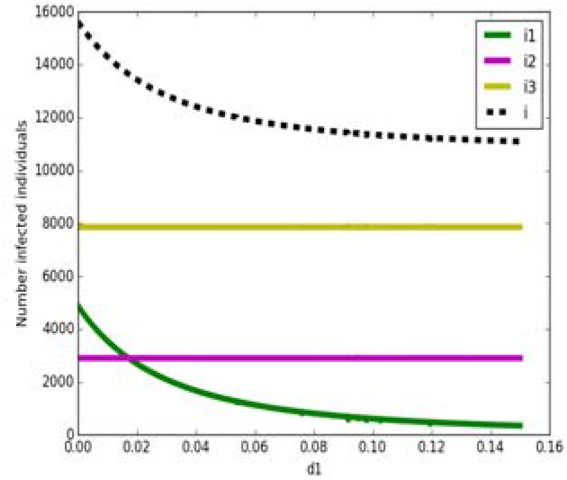

**Figure 4(i).**
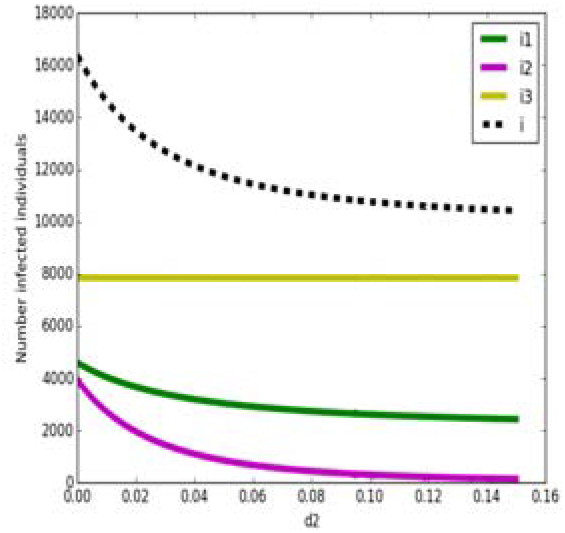

The following observations can be mentioned-

1. The total number of cases increases as b increases. On the other hand, the total number of cases decreases gradually as d_1_ and d_2_ increase.
2. For varying each parameter, the number of post reproductive infected people remains almost constant.
3. The number of reproductive infected individuals remains almost constant when bifurcation is performed with respect to b or d_1_ but decreases gradually when d_2_ is increased.
4. The number of infected pre reproductive population is sensitive to all the bifurcating parameters.
5. The black dotted curve resembles of the solid green curve not in magnitude but in sense. This implies that controlling the infection in the pre reproductive population would have a major impact on the total cases.

However, the total number of cases changes continuously and quite uniformly with the change of bifurcating parameters. Sudden increase or decrease is not observed within this range of the parameters.

### Section V: Two parameter bifurcation

Now we choose two parameters among b, d_1_ and d_2_ and vary them from 0.0 to 0.15 while keeping the other parameters fixed. Then we check the conditions c1 to c6 and then plot the points with the same color as mentioned in Section-III. Figure 5(a), (b), (c) are two parameter bifurcations of Nigeria; Figure 5(d), (e), (f) are two parameter bifurcations of USA and Figure 5(g), (h), (i) are two parameter bifurcations of Japan. The blue and sea green regions imply that pre reproductive population is mostly affected; Red and Olive-green regions show that the number of infected reproductive population is highest and the green and purple regions suggest that the post reproductive infected people is highest in number.

**Figure 5(a).**
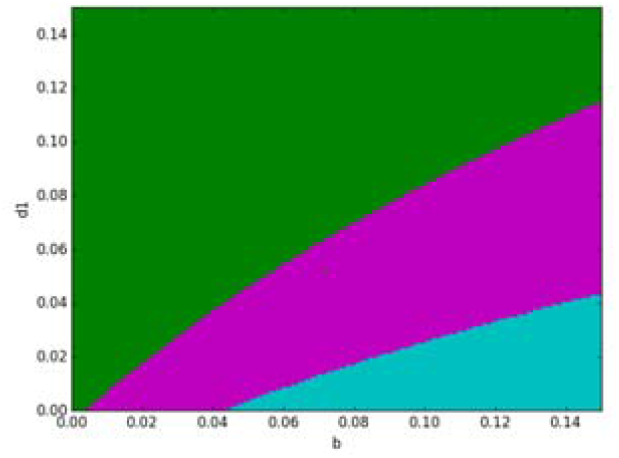

**Figure 5(b).**
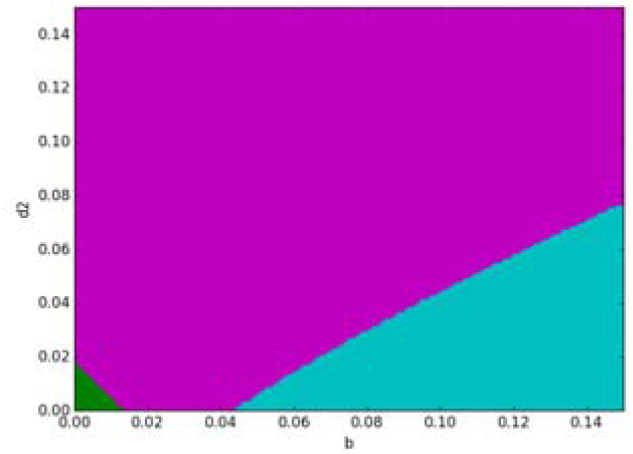

**Figure 5(c).**
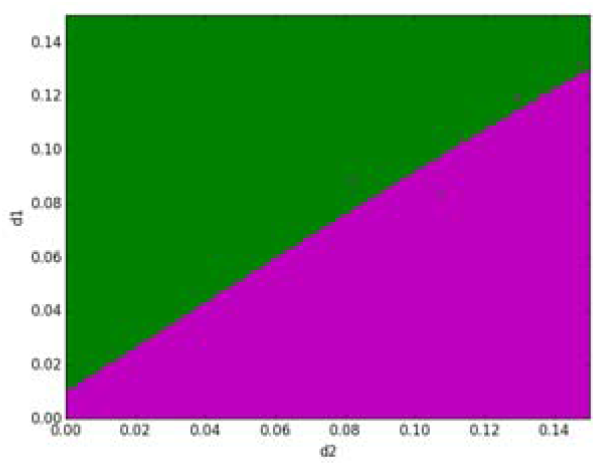

**Figure 5(d).**
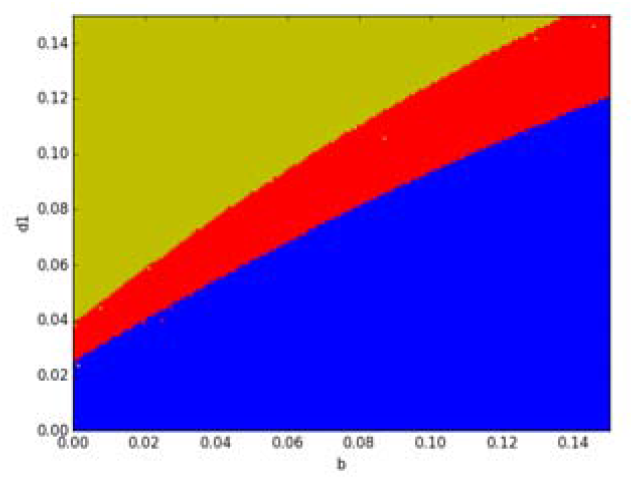

**Figure 5(e).**
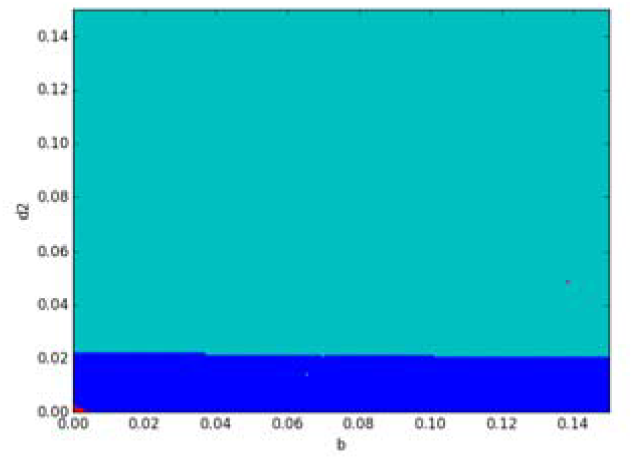

**Figure 5(f).**
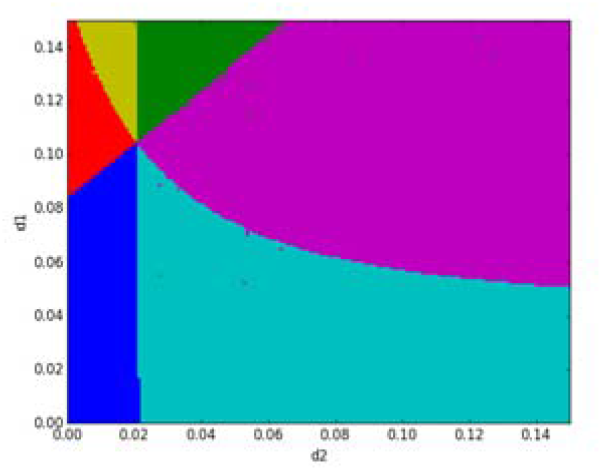

**Figure 5(g).**
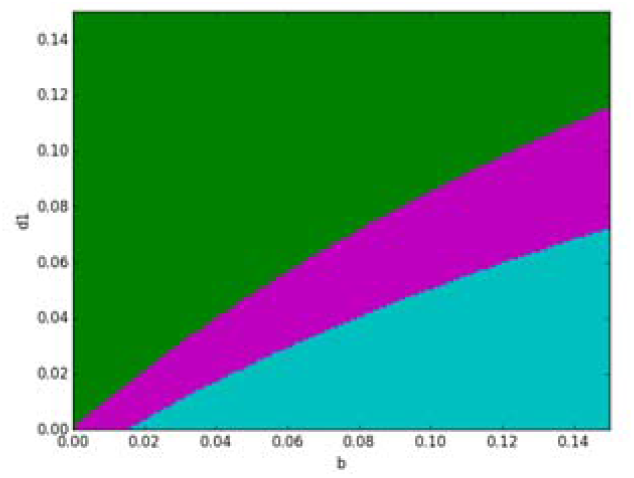

**Figure 5(h).**
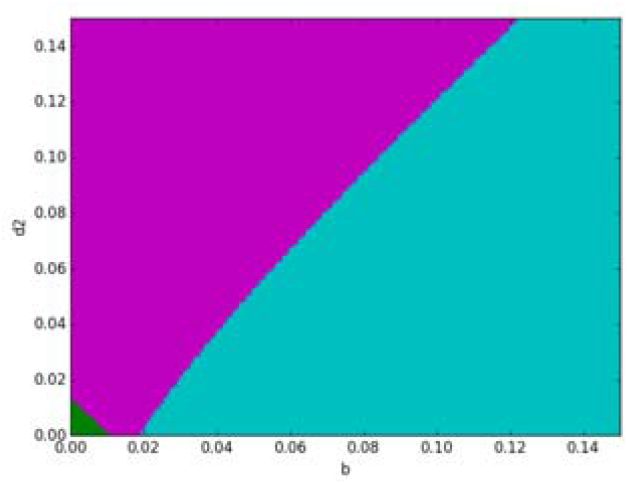

**Figure 5(i).**
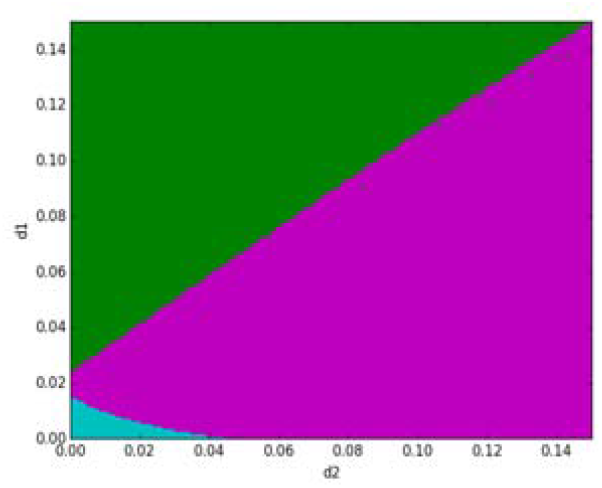

We see that if a country does not possess population pyramid of expanding type, the green and purple region is found for a broad range of values of bifurcating parameters. This implies that if the nation does not have expanding population pyramid, then the post reproductive individuals are at high risk. Blue and sea green regions that imply c1 and c2 respectively- are also found for a significant number of values of parameters.

### Section VI: Three Parameter Bifurcation

In this part we vary all the three parameters b, d_1_ and d_2_ from 0.0 to 0.15, then check the conditions c1 to c6 by studying the dynamics and finally plot different conditions with different colors as mentioned in Section III. This three-dimensional plot gives solid regions which are very difficult to understand but still we show them. For better understanding we also present pie diagrams which consist of the percentage of point (or to say equivalently the approximate percentage of volume of the solid regions) showing different conditions.

Figure 6(a) – (f) are the three parameter bifurcation diagrams of a country belonging to the population pyramid of expanding type (Nigeria). Figure 6(g) shows the percentages of points that show different conditions as mentioned earlier in Section III. The colors also indicate same conditions as stated in Section III.

**Figure 6(a).**
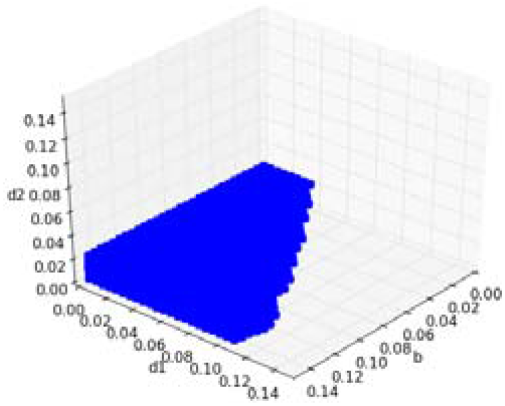

**Figure 6(b).**
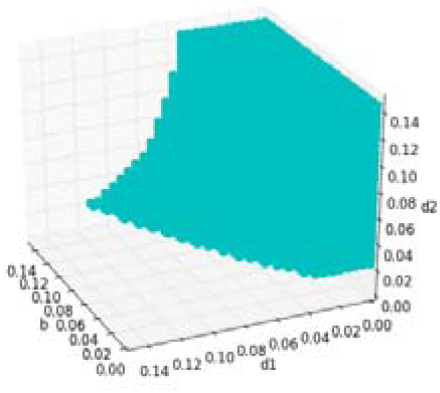

**Figure 6(c).**
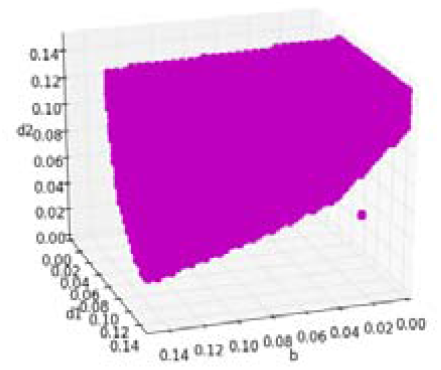

**Figure 6(d).**
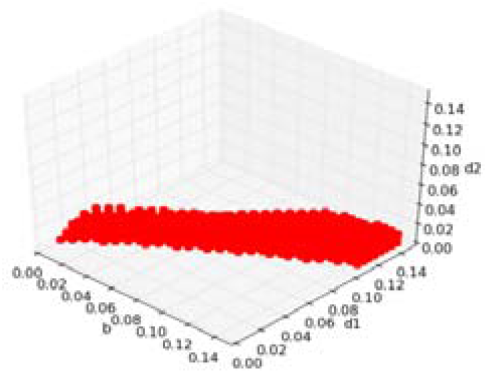

**Figure 6(e).**
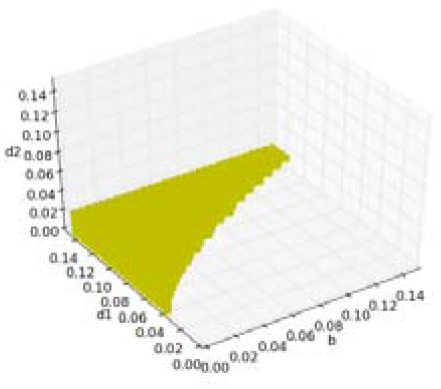

**Figure 6(f).**
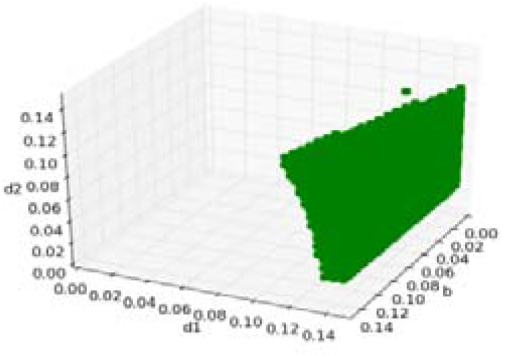

**Figure 6(g).**
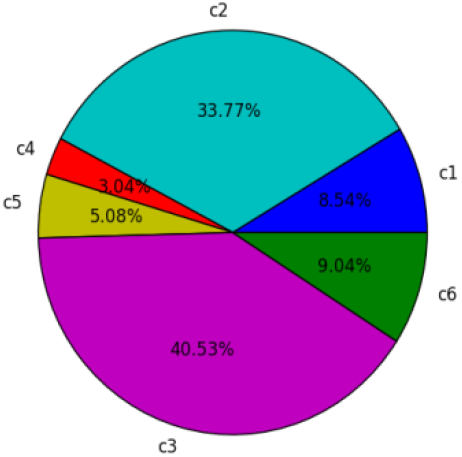

Next, we show the bifurcation diagrams of other two types of population pyramids. But since the blue, red and olive-green region disappears, we do not show them.

Figure 6(h) – (j) are the three parameter bifurcation diagrams of a country belonging to the population pyramid of stationary type (USA). Figure 6(k) shows the percentages of points that show different condition.

**Figure 6(h).**
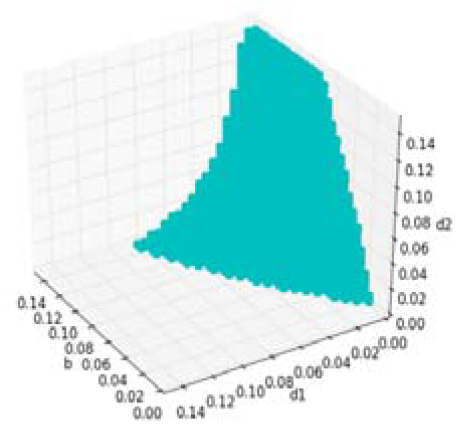

**Figure 6(i).**
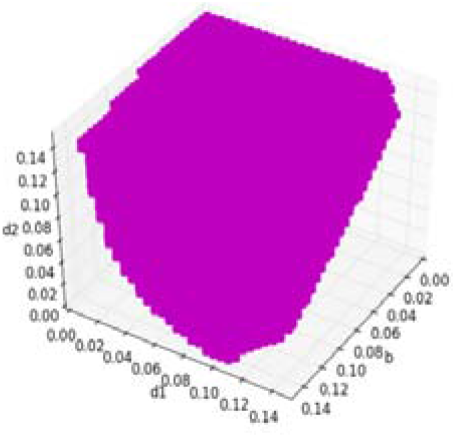

**Figure 6(j).**
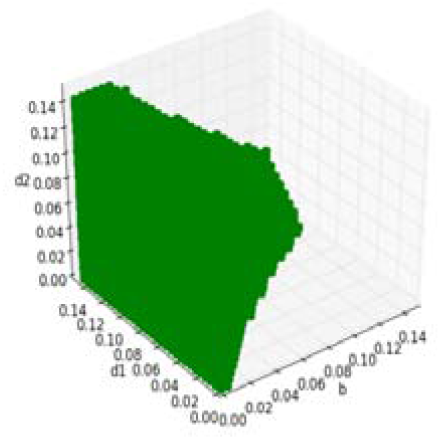

Figure 6(l) – (n) are the three parameter bifurcation diagrams of a country belonging to the population pyramid of contracting type (Japan). Figure 6(o) shows the percentages of points that show different conditions.

**Figure 6(k).**
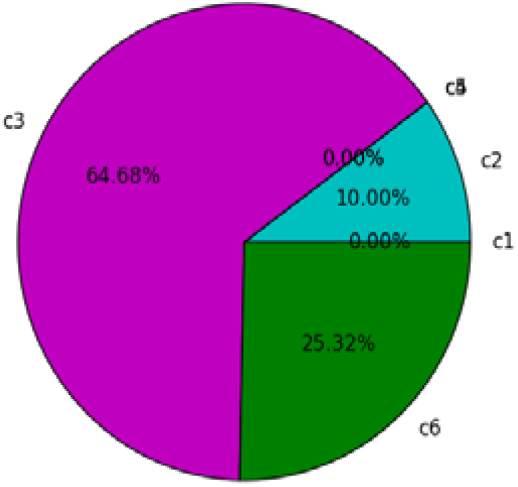

**Figure 6(l).**
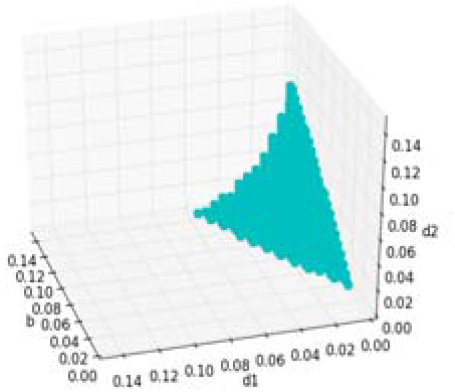

**Figure 6(m).**
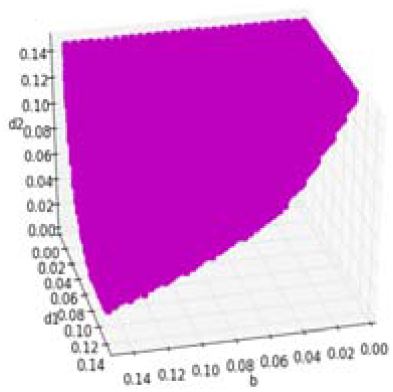

**Figure 6(n).**
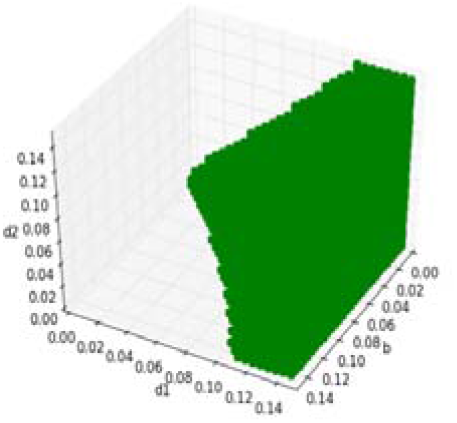

**Figure 6(o).**
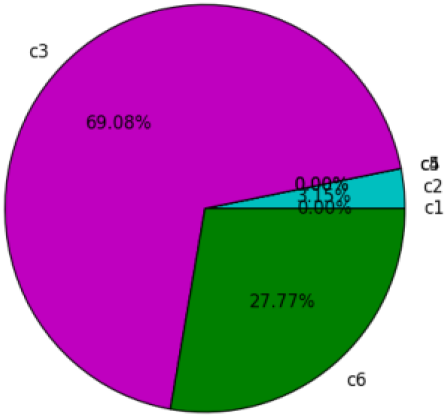

### Section VII: Dependence of total cases on Population Pyramid

Now we try to find conditions which refer to the reduction in the number of total cases. In Section I and IV we calculated the total cases for three different countries and for different values of chosen bifurcating parameters. There we found that four cases may arise- (a) the total number of cases is less than 5000; (b) the total number of cases is more than 5000 and less than 10000; (c) the total number of cases is more than 10000 but less than 15000; (d) the total number of cases is more than 15000. In figure 7(a) we perform initial value sensitivity in the similar fashion as described in Section II and divide the plane x +y +z =1000 into four distinct parts denoting the above mentioned four cases. Let the green color represent first case; the sea green, olive green and red regions denote second, third and fourth cases respectively though we do not find any red regions for the chosen values of b, d_1_ and d_2_. It is observed that the total number of cases is low when pre reproductive population is high and reproductive and post reproductive population is low.

**Figure 7(a).**
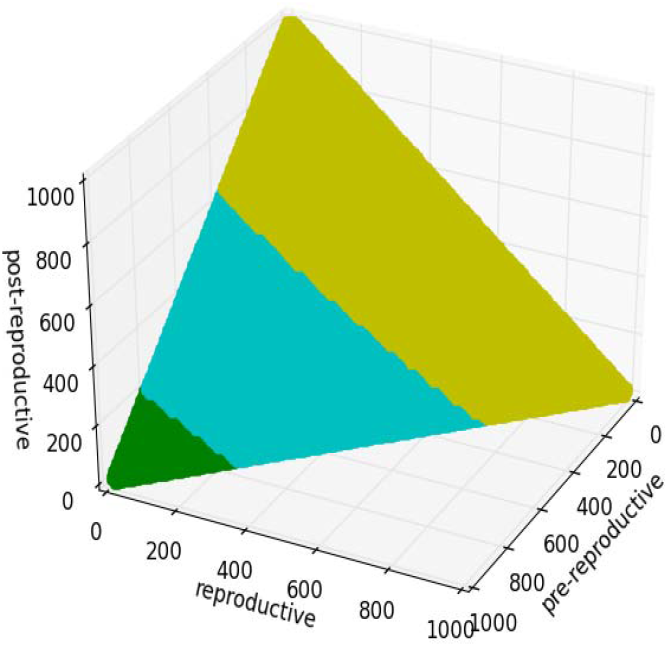

The total number of cases is high when pre reproductive population is low and reproductive and post reproductive population is high for a fixed set of values of parameters.

So now we vary all of b, d_1_ and d_2_ from 0.0 to 0.15 for three countries mentioned in Table 3 and check the above cases. We also plotted them but we did not show them here. A curious reader may find it in the supplementary file. However, we show the pie charts of the percentage of parameter values that show the cases stated above. Further we state some important results.

1. The green region is found only in case of Nigeria which implies that if the population pyramid is of the type stationary or contracting then it is quite obvious that the total cases will be much more than the expanding population pyramid.
2. Lower values of d_1_ and d_2_ increases the total number of cases.

Figure 7(b), (c) and (d) are the pie diagrams of Nigeria, USA and Japan respectively.

**Figure 7(b).**
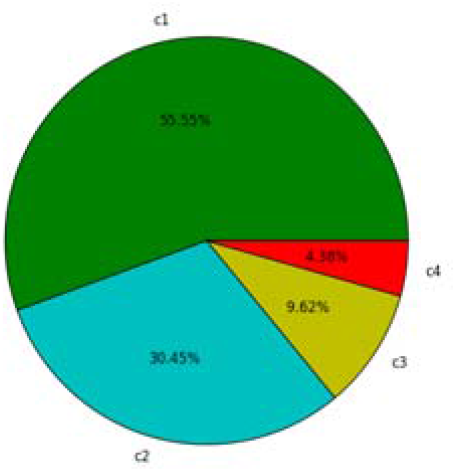

**Figure 7(c).**
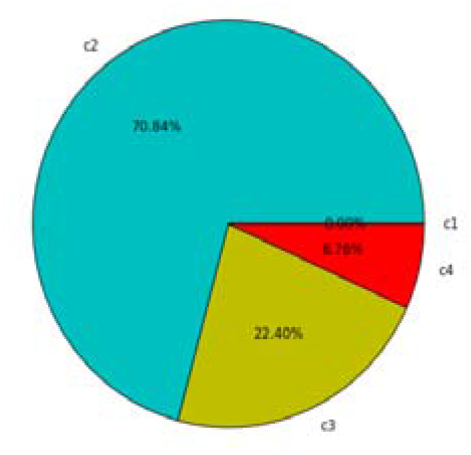

**Figure 7(d).**
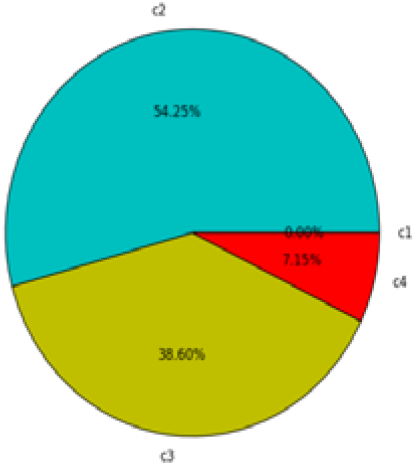

## Results

1. From TABLE-4 of Section II we found that for the countries belonging to Population Pyramid of expanding type, the non-vaccinated age group (i.e., the pre reproductive age group) consists a major portion (i.e., about three fourth) of the total infected people. This result once again supports the importance of vaccination (Moghadas, 2020). Researchers showed that vaccination can reduce the total cases almost half (COVID research: a year of scientific milestones) of the prediction.
2. In Section III we observed that the purple and green region take a major portion (almost 57.14%) of the plane (Figure 3(a)). The purple and green portions suggest c3 and c6 respectively which implies the post reproductive age group is affected mostly. Thus, it can be concluded that the post reproductive age group has a tendency to be infected more than other age groups for a huge number of the initial populations.
3. From Figure 3(a) and 3(b) we also find that the blue and sea green region have a high portion. This implies that the pre reproductive population is also affected mostly for a significant number of initial populations.
4. Further it is also observed from Figure 3(a) and 3(b) that the reproductive population is affected mostly only when they consist almost the whole population.
5. From Figure 4(a)-(i) it is clear that for each country there is a constant number of infected post reproductive people independent of different values of b, d_1_ or d_2_.
6. The age group that will be infected most, depends highly on the values of parameters.
7. It is clear that the post reproductive age group is more prone to infection for the countries having population pyramid of stationary or contracting type. So special precautions are required to be taken to save that age group from infection in those countries. Their vaccination should be given the most priority.
8. The pre reproductive population is also at high risk for several values of parameters especially for the countries with expanding population pyramid.
9. The total number of cases also depends on the population pyramid. If the pre reproductive group is high in number, the total case is comparatively low. As the number of individuals in reproductive and post reproductive group increases and pre reproductive group decreases, the total cases increase.
10. From Figure 7(b) to (d) the percentage of red region increases that implies highest number of cases occur for more values of parameters if the population pyramid of the country is of the type contracting instead of expanding.
11. If the population pyramid is of the types stationary or contracting, then very low number of cases (i.e., the green region) is not found for any value of the parameters. Thus, it is quite expected that in those countries, the number of cases will be more compared to the countries with population pyramid of expanding type.

**Table 4:** Some important observations based on the numerical simulation.

| **Country** | **Total number of infected populations** | **Total number of infected pre reproductive population** | **Total number of infected reproductive population** | **Total number of infected post reproductive population** | **Percentage of infected pre reproductive population** | **Percentage of infected reproductive population** | **Percentage of infected post reproductive population** |
| --- | --- | --- | --- | --- | --- | --- | --- |
| Nigeria | 18415 | 13567 | 2924 | 1924 | 73.67% | 15.88% | 10.45% |
| USA | 16236 | 6887 | 3446 | 5903 | 42.42% | 21.22% | 36.36% |
| Japan | 14874 | 4125 | 2896 | 7853 | 27.74% | 19.47% | 52.79% |

## Discussion

Our study contained a details analysis of disease dynamics in countries with different types of population pyramids through an age-based population model and it reveals several important conclusions. Firstly, in the countries having population pyramid of the type stationary or contracting, the number of cases is observed to be high unlike the countries having population pyramid of expanding type where the total case is observed to be low according to our experiment. So, if a state does not have expanding population pyramid, needs highly developed health-infrastructures and should follow the preventive measures strictly with high consciousness. In the global vaccination program (Mathieu et al., 2021; Greenwood, 2014), such countries deserve high attention.

Secondly, the Covid-19 pandemic has a tendency to attack the post reproductive age group (in spite of vaccination) in the countries that have stationary or contracting population pyramid and the pre reproductive age group in the countries that have expanding population pyramid. The reproductive group, consisting mainly young generation, is comparatively at low risk in all the countries. So instead of strict lockdown, an age specific partially strict lockdown is more preferable biologically and economically as well. Thirdly, from the initial value sensitivity we can conclude that the Covid-19 is transmitted mostly in post reproductive age group unless its initial population is too low. Another important fact is that for a particular country, the total number of infected post reproductive population does not depend on b, d_1_ or d_2_.

## Conclusion

This paper focuses on the numerical results and their significances. So, we did not discuss the theoretic analysis, basic reproduction number and next generation matrix. For numerical simulation, it was assumed that 1%, 2% and 3% people of three age groups are infected initially. This is an over estimation. But we assumed it so that we obtained results that are easy to understand.

The three countries chosen here i.e., Nigeria, USA and Japan do not deserve any special attention. They were selected in a completely arbitrary manner just to have an idea about their initial populations of different age groups and to determine the values of several demographic parameters such as birth rate, death rate etc. By this choice of the countries, we cover different socio-economic and geographic perspectives. It is useless to try to match the number of cases with actual data. But the obtained results hold good for not only those three countries but also for several countries having similar population structure.

## Data Availability

All data produced in the present work are contained in the manuscript

## Authors’ Contributions

Conceptualization, Supervision: TS; Data curation, Formal analysis, Writing - original draft: AB; Investigation, Methodology, Writing - review and editing: TS, AB.

## Acknowledgement

We are really thankful to Dr. Uttam Ghosh & Mr. Aloke Saha for their kind advice.

